# A multiagent coronavirus model with territorial vulnerability parameters

**DOI:** 10.1101/2020.10.25.20218735

**Authors:** Patrícia Magalhães, José Paulo Guedes Pinto, Diana Maritza Segura Angel

## Abstract

We developed a simple and user-friendly simulator called MD Corona that is based on a multiagent model and describes the transmission dynamics of coronavirus for a given location considering three setting parameters: population density, social-isolation rate, and effective transmission probability. The latter is represented by the Coronavirus Protection Index (CPI) - a measurement of a given territory’s vulnerability to the coronavirus that includes characteristics of the health system and socioeconomic development as well as infrastructure. The dynamic model also relies on other real epidemiological parameters. The model is calibrated by using immunity surveys and provides accurate predictions and indications of the different spread dynamic mechanisms. Our simulation studies clearly demonstrate the existence of multiple epidemic curves in the same city due to different vulnerabilities to the virus across regions. And it elucidates the phenomenon of the epidemic slowing despite a reduction in social-distancing policies, understood as a consequence of “local protection bubbles.” The simulator can be used for scientific outreach purposes, bringing science closer to the general public in order to raise awareness and increase engagement about the effectiveness of social distancing in reducing the transmissibility of the virus, but also to support effective actions to mitigate the spread of the virus.

## 1- Introduction

Modeling SARS-CoV2 is crucial to understanding the dynamics of the virus’s transmission. Models are useful for describing spatial and temporal patterns of disease prevalence, exploring the effectiveness of confinement measures to reduce the incidence of infection, and helping predict the potential resurgence of new waves of infection. But to understand the complexity of the virus’s epidemic curves in Latin America, particularly in a continent-sized country like Brazil, we have to consider the diverse socioeconomic structure of the territory in which it spreads, because the narrative based on a single uniform contagion curve hides the inequalities between different populations and territories.

Our model is an agent-based complexity model, which simulates the spread of the coronavirus in various territories. This allows users to gauge local vulnerability to the virus by linking the probability of transmission between agents with the Coronavirus Protection Index (CPI). Developed by our research group (Ação Covid-19, 2020a), the CPI accounts for characteristics of the health system, socioeconomic development and territorial infrastructure indicators for many Brazilian districts and towns that have a direct influence on the virus’s spread.

We have used MD Corona to develop numerous studies on vulnerability to Covid-19 in different Brazilian cities: Fortaleza, São Paulo, Rio de Janeiro and Curitiba (Ação Covid-19, 2020b; Ação Covid-19, 2020c; Ação Covid-19, 2020d; Ação Covid-19, 2020e). Using the model, we demonstrated that there are multiple epidemic curves in a single city. Once the model is calibrated it can make very accurate predictions. The most important prediction relates to the phenomenon observed in some cities where the number of new infections decreased despite the reduction of social-distancing policies. Our model showed that this can be understood as a consequence of local bubbles of protection formed in city sub-environments, together with the exhaustion of infection networks (Guedes Pinto et al., 2020).

Our paper is organized as follows: In section 2, we review the main characteristics of different modeling approaches currently being used to describe coronavirus epidemic curves. In section 3, we present our model in detail, as well as the calibration procedure that allows us to make predictions for specific territories. In Section 4, we show some examples of the power of the model in different case studies that compare the dispersion of Covid-19 in different vulnerable territories. Finally, in section 5, we conclude by highlighting the possible uses of the model, some recommendations, and perspectives for the future.

## 2. SARS-CoV-2 Models

Modeling the Coronavirus epidemic curve is a complex task, as one needs to consider a wide number of nonlinear parameters such as the implementation of non-pharmaceutical interventions (NPI) as well as local social and territorial characteristics. In addition to our multiagent model approach, there are also SIR or SEIR model families, based on multiple differential equations, and statistical models. Obviously there are also combinations of all of these methods.

The *SIR/SIRS and SEIRS/SEIR* family of models refers to the transitions between the different epidemiologic states that are attributed to individuals in each model – that is, susceptible (S), infectious (I), recovered (R), and exposed (E). These models can become even more complex by introducing new epidemiologic states and different parameters, such as the use of NPI. Examples of the use of this model are presented by Giordano et al. (2020), Penn Medicine (2020), Yeghikyan (2020), and Medeiros et al. (2020).

The main characteristic of these models is their extreme sensitivity to small changes in parameters. To fix R0 - the initial disease reproduction rate - and other parameters, these models must use available data on the number of infections and death over time. Examples of the use of this model in interactive simulations are presented by Alves et al. (2020), Neiva et al. (2020), Flaxman et al. (2020), Li et al. (2020), Goh (2020), and Hill (2020). Although these models can be very precise for a given set of data, they can be very difficult to understand and to manipulate without scientific knowledge of the field.

*Statistical models*, meanwhile, are probability distributions that are able to describe different patterns. The target of statistical inference is to identify which specific functions and parameters can reproduce the available data and make predictions about future sets. The main statistical models are probabilistic and multivariable models. The former is related to frequentist and Bayesian theories, whereas multivariable models focus on assessing the relationship between a single dependent variable and multiple independent variables (Stokes, Davis and Koch, 2020). An example of this model is presented by Allenbach (2020).

Statistical models do not simulate the virus’s spread based on epidemiological assumptions and equations, but by fitting a curve to prior data. Therefore, they cannot adjust to socioeconomic and mobility parameters or to territorial or population characteristics. Their main weaknesses are that they do not provide accurate results at the beginning of a pandemic and their strong dependency on actual data. Particularly in Brazil, this can be a major problem due to widespread underreporting of cases and inefficient testing practices (Covid-19 Brasil, 2020b). The authors of some statistical studies say that it is hard to predict where we are in the pandemic curve without precise information about the real number of infected and dead people (Wynants, 2020).

Finally, in *agent-based models* (Ajelli et al., 2010; Venkatramanan et al., 2017) the phenomenon is described by the successive interactions between individual objects with particular properties and actions (Wilensky and Rand, 2009). Agent-based representations have particular advantages, such as the ease of graphically understanding the interactions between agents due to simple rules for their movement, as opposed to differential equation models that are developed from mathematical constructs. The increasingly widespread adoption of agent models provides some benefits to the understanding of hard domains in a more accessible way (Wilensky and Rand, 2009). Unlike SIR-based and statistical-model approaches, the multiagent model (Ajelli, 2010; Venkatramanan, 2017) does not depend on pandemic data (cases, deaths, and recoveries, for instance) to make predictions about the epidemic curve. However, like SRI-based approaches, the results of multiagent modeling of the virus’s spread depends on the initial conditions, such as mobility, geographic conditions, and population characteristics (Yeghikyan, 2020).

Our “MD Corona” model, subscribing to the latter model described above, simulates the dispersion of the coronavirus based on complex multiagent variables in the NetLogo environment (Wilensky, 1999). One unique feature of our model is the inclusion of socioeconomic, infrastructure, and health conditions through an effective transmission probability, but the details will be described in the next sections of this article.

There are other examples of this type of approach, such as Romer (2020), Steves (2020), Alves et al. (2020), Neiva et al. (2020) and Scabini et al. (2020), the latter being an example of a sophisticated multi-layer model produced by a Brazilian research group^1^.

Our (open-source code) simulator, in contrast to these precise and complicated models, was designed for users without a scientific, mathematical, or computational background. We also provide videos and user guides on our research group’s website (https://acaocovid19-homolog.web.app/dash).

## 3. MD Corona dispersion model

The main aim of the “MD Corona” model is to provide a simple tool for users to simulate the epidemic curve of SARS-CoV2 in neighborhoods and communities connected to large urban centers with different and distinct vulnerabilities.

It is inspired by the original virus model (Wilensky, 1998) present in the NetLogo free software library (Wilensky, 1999) supported by the work of Yorke et al. (1979), which suggests a number of factors that could influence the virus’s survival and transmission within populations.

In our model, individuals are agents moving randomly around a 41 x 41 grid. The agents can be displaced anywhere on this grid. The virus’s transmission then depends on whether the interaction between two or more agents (infected, immune, or susceptible to infection by the virus) in a von Neumann neighborhood^2^ can result in one agent infecting the other(s).

The dynamics of the coronavirus’ spread are driven by some epidemiological constants such as i) the virus-transmission period and ii) the immunity period; but also by parameters that we can vary to describe different scenarios, such as iii) the number of agents on the grid, iv) the initial number of infected agents, v) the probability of the virus being transmitted between agents, and vi) the practice of social distancing. The vii) recovery rate is also an epidemiological constant, but only affects mortality from the virus, not the dynamics of its spread.

Below, we will define each of these seven factors, justifying the choice of values that we assumed based on the most recent medical literature available. Because of the newness of this pandemic, one must note that those values are constantly changing in response to results from new studies. Users can modify all of these variables, since the code is open source.

The i) *virus-transmission period* varies a lot in the medical literature and depends on the disease’s severity, e.g., for patients with mild symptoms this value can range from 7 to 12 days (Ferguson et al., 2020, Liu et al., 2020, Cao et al., 2020). Due to the wide-ranging variation for this period, and the WHO (2020) recommending an isolation period of 14 days, we adopted the WHO value and added a four-day incubation period, therefore establishing the transmission period at 18 days, since there are reports of virus transmission in this period (ECDPC, 2020; CDC, 2020; Pan et al., 2020; Qian et al., 2020; Zou et al., 2020 and Zaki, 2020).

Many issues arise when we try to establish an average ii) *immunity period* to SARS-CoV2. A few studies state that the antibodies’ response to infection varies depending on the duration of the infection and the severity of the disease (Luchsinger et al., 2020; Seow et al., 2020), and because of that they don’t provide an average length of time for patient immunity. Other works (Wu et al., 2007; Wei Liu et al., 2006; Edridge et al., 2020; Cao et al., 2007; Kellam & Barclay, 2020) report that antibody responses to other human coronaviruses (SARS-CoV, MERS, alpha and beta coronaviruses) wane over time, varying from 12 weeks to 34 months.

Based on the abovementioned literature, and due to the newness of the disease, we chose to consider the immunity period as the time that the pandemic has lasted so far, thereby setting the agent’s immunity period at 180 days. It is important to add that some research, such as Serrano (2020), affirms that mutations in the virus strains mean that a relatively long time will be needed to produce a vaccine.

The iii) *number of agents in the grid* (or *population density*) is an important parameter in the model, since it affects the frequency of contact between agents in the grid and consequently the probability of the virus’s transmission between infected and healthy people. To make the simulator easier to use, in the version available on our website we converted the variable ‘number of people’ in the grid into a ‘demographic density’ variable (set with sliders), which allows users to easily adapt the simulator to a territory of their choice. The coefficient that converts ‘number of people’ to ‘demographic density’ is defined throughout a calibration method discussed below.

The iv) *number of agents initially infected* was fixed at one person regardless of the total number of people in the grid. The model also allows a periodic reintroduction of a new infected agent (the frequency of which can vary, depending on the scenario we wish to simulate). This permits the appearance of new infection waves and for the system to remain open, which is consistent with the reality of the virus’s circulation across different territories.

The v) *virus’s transmission* between agents depends on a number of non-pharmaceutical interventions such as the use of masks, hygiene procedures, and social-distancing measures. But it is also affected by social conditions and particularities of each territory, such as the existence of basic sanitation, the average number of people per house, if families are capable of implementing social distancing, etc. This mixture of territorial, health, and social factors are not easy to quantify. We parameterize them in the simulator through an effective transmission probability, which is directly related to an index – either the Human Development Index (HDI) or the Coronavirus Protection Index (CPI), the latter being an innovation that came along with this model and that was also developed by our research group (Ação Covid-19, 2020a).

The HDI has the advantage of being universal, but it serves as a very poor measurement of different territories’ vulnerability to coronavirus. The CPI, on the other hand, was based on the Surroundings Index (SI) methodology (Ranieri & Begalli, 2016) and includes other aspects, such as characteristics of the health system, socioeconomic development and infrastructure indicators.

We classified a large number of districts and cities in Brazil by using CPI, and this resulted in a more coherent evaluation of vulnerability in a specific territory. Both HDI and CPI are divided into 5 levels: very high, high, medium, low, and very low (see Figure 1). Fine tuning the effective probability rate and the HDI/CPI scale is carried out through calibration.

**Figure 1.**
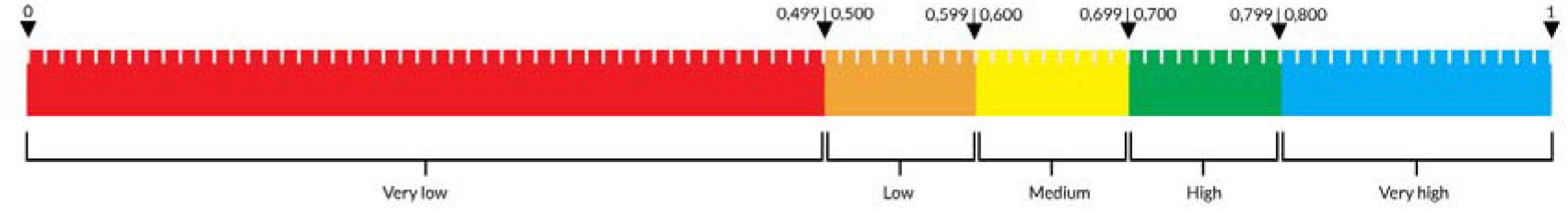
Ranges for the different levels of HDI and CPI.

Another important innovation of this model is the inclusion of the vi) *social-distancing rate* as a dynamic parameter in the model. This slows down the spread of the virus by not allowing a portion of the agents to move, and the rate can vary while the simulation is running to describe changes in social distancing over time.

Finally, the vii) *infection-fatality rate* (IFR) determines how deadly a disease is. This is calculated as the proportion between the number of infected patients and the number of deaths, including asymptomatic and undiagnosed infections. The IFR also depends on local health conditions (hospital access and occupancy rates) and age factors. There are a few estimates for infection fatality rates in Brazil that use the total number of deaths and seroprevalence surveys. Mallapaty (2020) used a sample of 25,025 participants from all 27 Brazilian states (Hallal, 2020), suggesting a rate of 1% for the IFR. However, a more accurate seroprevalence survey carried out only in the city of São Paulo, with a sample of 5,416 participants, found an IFR of 0.7% (G1, 2020). The same methodology was also used to compute an IFR of 1% for Spain (Ministry of Health, 2020), 0.7% for France (Salje, 2020), and 0.66% for China (Verity, et al., 2010). Considering that we are interested in simulating the spread of the virus in urban regions, we chose to use the São Paulo IFR measurement of 0.7% as a constant in our model for all the different territories.

### 3.1 How does the model work?

To begin the simulation, the user should define the number of people (agents) in the grid and the effective transmission probability by setting the population density (through a slider) and the HDI/CPI (through a chooser) after consulting a table we provided in the simulator that displays information for neighborhoods or districts in several Brazilian cities. In Figure 2, we can see that the remaining parameter is ‘social distancing,’ where the user can choose on the slider the proportion (from 0% to 100%) of agents that will randomly cease to move within the grid. That rate can be changed while the simulation is running.

**Figure 2.**
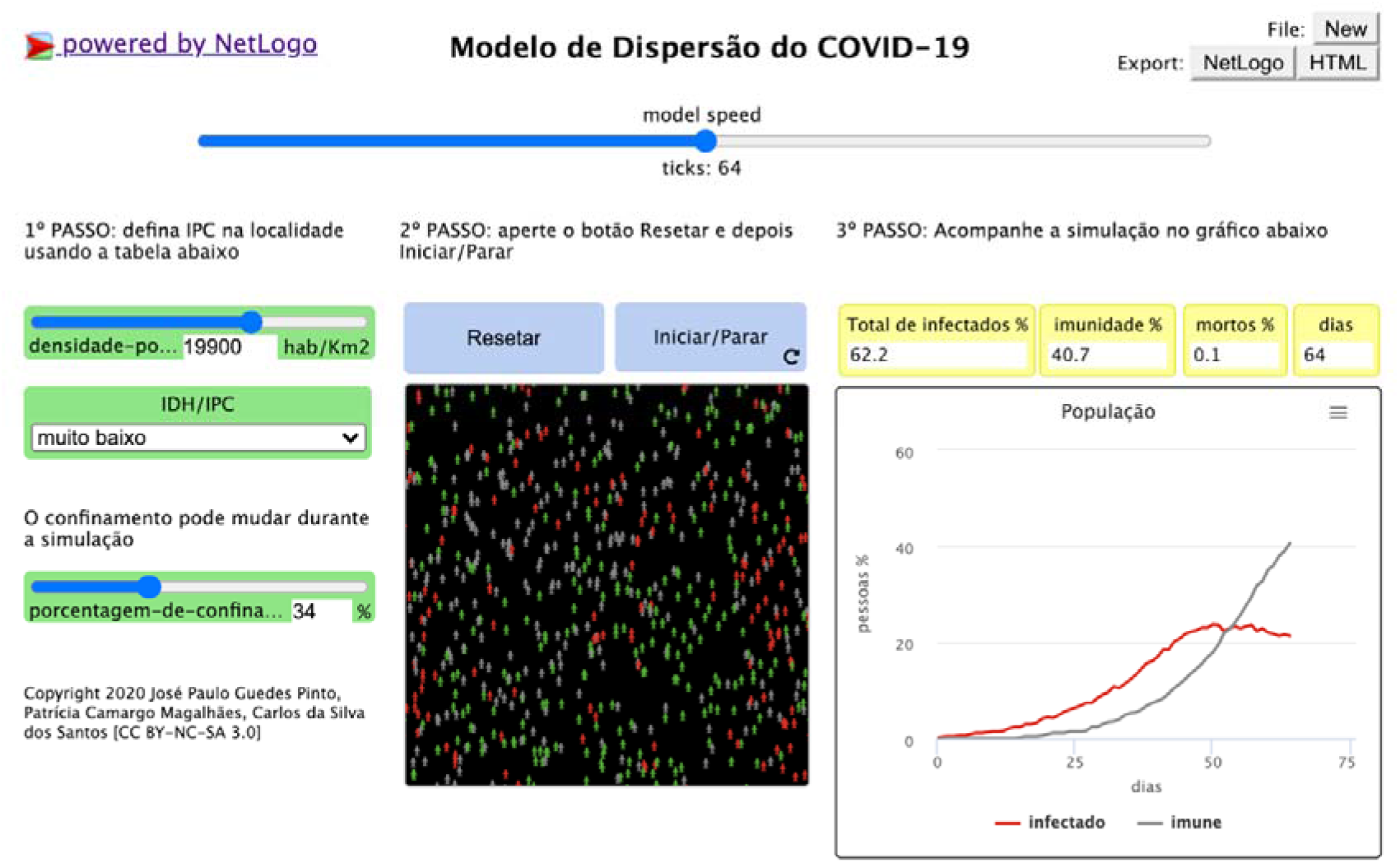
MD Corona Simulator version 3.0 on our website.

The model’s time scale was set in days, and each round is equal to one day. Agents that move randomly in this environment are classified into one of three states: healthy agent (green), infected agent (red), or immune agent (gray), as can be seen in Figure 2. When running the simulation (clicking ‘reset’ and then ‘start’), the virus’s transmission is the outcome of the meeting of at least two people on the grid and the actual likelihood of infection.

The number of people infected appears in a graph, along with the number of people who have become immune (immunity curve). A counter shows the number of simulation days and the percentage of infected, immune, and dead within the population (the simulation speed can be defined by the user on a slider).

People can die from virus infection or due to age (75 years - the mean life expectancy in Brazil). When the population falls below the “maximum capacity” of the environment (set at 1,000 people), healthy people can produce healthy descendants (but susceptible to infection).

The “MD Corona” model generates stochastic processes, sensible to small changes in the initial conditions randomly determined by the simulator each time (the relative position of infected and immobilized individuals). We account for this fluctuation by running at least 100 simulations for each scenario with a Python support program we developed, which allows us to extract and analyze the average results. A more complete stochastic analysis could investigate the different trends in the simulation results, identifying the behavioral change threshold positions, i.e., where the curve’s inflection points are. This will be done in an upcoming publication.

### 3.2 Model Calibration

In our first version of the model, the main feedback we received was about the need for some way to set parameters to describe realistic environments. Users wanted to know what population size they should use to simulate their territory. Therefore, we developed a technique to calibrate the model and connect the number of agents in the grid to a specific population density, and the effective transmission probability to the respective HDI/CPI scales by using seroprevalence surveys and the trajectory of social isolation for each specific place.

It is important to stress that in our model the epidemic curve included symptomatic and asymptomatic patients. This is why in order to calibrate the model it is necessary to use the results of studies that test immunity to the virus, i.e., seroprevalence in random populations. These tests detect the presence of Immunoglobulin G (IgG) antibodies produced by people who have been infected with the SARS-CoV-2 virus for at least 20 days.

In terms of population density on the grid (number of agents/1,681 points on the grid), the choice of the number of people on the grid representing the calibration place determines the range of territorial densities we can cover within the simulator. In all cases, however, the calibration methodology is exactly the same: users plug in the known history of social-distancing rates in the simulator and toggle the effective transmission probability and the number of people in the grid to match results from seroprevalence surveys on a given date, so that time-lapse simulations arrive at the same percentage of infected people as reported in survey data.

In the case of the city of São Paulo, for example, applying the known history of social-isolation rates (Governo do estado de São Paulo, 2020) given in Table 1, we set the number of agents in the grid to 369 and the effective transmission probability at 40%, corresponding to a population density of 8,054.7 inhab./km^2^ and the ‘high’ level (0.79) on the CPI scale, respectively.

**Table 1.** History of social-distancing rates for the city of São Paulo used in the calibration of “MD Corona”

| Date | Description | Social-distancing rate | Number of days |
| --- | --- | --- | --- |
| Feb 25 – Mar 17 | 1st case and self-imposed social distancing | 27% | 21 |
| Mar 18 – Mar 21 | official social constraints | 43% | 4 |
| Mar 22 – Apr 12 | decline in social distancing (Bolsonaro effect) | 58% | 22 |
| Apr 13 – May 3 | further decline (Bolsonaro effect) | 53% | 21 |
| May 4 – May 31 | announcement of São Paulo plan | 51% | 28 |
| June 1 – June 22 | São Paulo plan - seroprevalence survey 9.5% | 48% | 22 |
| June 23 – July 12 | orange and yellow phase | 46% | 20 |
| total days simulated |  |  | 138 |

Source: São Paulo State Government (2020)

With these parameters set, the simulation results (Figure 3) show that 10.87% of the São Paulo population were infected, on average (the colored curves represents each simulation), and an average mortality rate of 0.08% (or 0.75% of the total infected) on June 22, 118 days after the first case. This is compatible, as expected, with the immunity survey that reported 9.5% (with a 1.7% error interval) of São Paulo residents infected by Covid-19 on the same date (G1, 2020).

**Figure 3.**
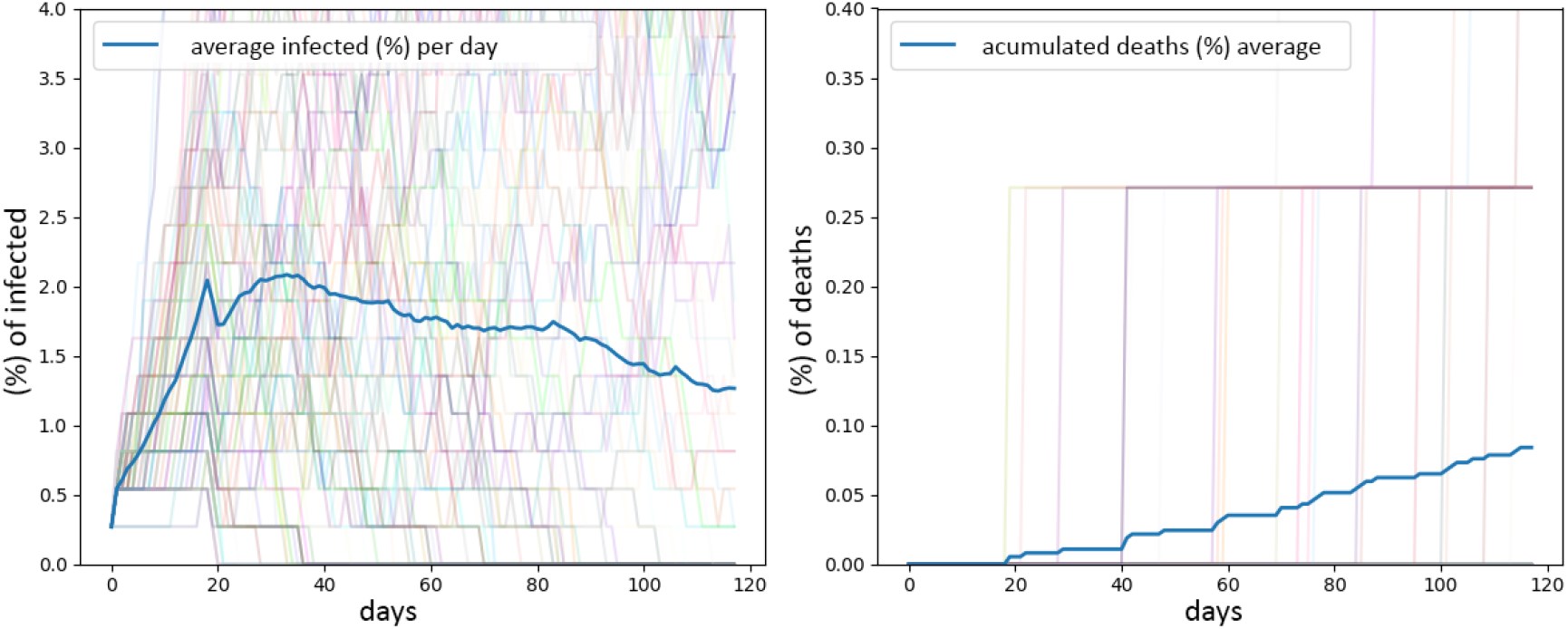
Results for 100 simulations of MD Corona model with the history of confinement given in table 1: (left) the percentage of infected people (daily), totaling 10.87% after 118 simulated days; (right) percentage of deaths, totaling 0.08% after 118 simulated days. The blue curve is the average curve for all the simulations.

By calibrating the model for São Paulo, we established the city’s ‘high’ CPI level with a 40% probability of transmission. Using the same calibration interval, we applied it to New York City (10,947 inhabitants/km^2^ and ‘very high’ HDI) – where the most recent immunity survey reported 19.9% of people infected as of May 2 (Governor Andrew M. Cuomo’s Press Office, 2020) – by setting the transmission probability at 39%. Therefore, by carrying out this calibration, the model converted each element on the scale (HDI or CPI) into respective transmission probabilities of 39%, 40%, 42%, 44%, and 46% for “very high,” “high,” “medium,” “low,” and “very low” HDI/CPI levels.

The calibration procedure established a rule for converting demographic densities into the number of people that should be entered into the simulator:

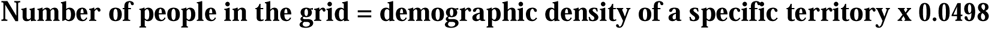

This particular calibration defines a maximum (20,080 inhab./km^2^) and minimum (3,010 inhab./km^2^) range for population density in the simulation, considering that the number of agents in the grid can vary from 150 to 1,000. For densities greater than the maximum, we recommend that users set the slider to the maximum level, as transmission dynamics will be similar.

The ease of changing MD Corona parameters allows us to implement different calibrations depending on the specific characteristics of the territory we want to study and the availability of immunity surveys and social isolation data. The very first calibration we performed in late April used New York City’s first immunity survey, one of the few available at the time, which reported immunity rates of 20% on April 22 (NY Times, 2020). More recently, in a study that included very high-density slums in Rio de Janeiro (Ação Covid-19, 2020f) we used results from a study in the city with 3,210 samples in six neighborhoods – including Cidade de Deus, a very dense slum with 28,684 inhabitants per square kilometer, where data showed that 28% of the population had been infected with Covid-19 (Ação Covid-19, 2020f).

## 4. The Model as a tool for understanding the unequal spread of Covid-19

From the very beginning of its development, MD Corona was meant to serve educational outreach and research purposes, but we have always considered its potential to be used, albeit cautiously, as a tool to help decision-making by public and private planners, as well as community leaders, social movements, and other civil society actors that could play a relevant role in this pandemic.

So far we have used MD Corona to develop numerous studies on vulnerability to Covid-19 in different Brazilian cities: Fortaleza, São Paulo, Rio de Janeiro, and Curitiba. With the CPI calculated for unequal neighborhoods in terms of socioeconomic development, we demonstrated the existence of multiple epidemic curves within the same city.

Once it is calibrated, there are multiple explorations one can develop with MD Corona: using the dynamics of confinement to tell a story about the virus in that environment and make very accurate predictions; studying the social-confinement rates needed to not overwhelm the health system and instead flatten the epidemic curve; or using the history of social isolation and immunity survey data to evaluate the effectiveness of NPI implemented in a specific territory. Also, the model reveals important features of the dynamics of the epidemic outbreak. Below we show examples of potential uses for the Model through some studies we have performed so far.

### 4.1. Different epidemic curves across districts of major Brazilian cities

We studied the epidemic curve of the virus in different districts of big cities that have distinct Covid-19 protection indexes. In Rio de Janeiro, we looked at (Ação Covid-19, 2020d) the famous Copacabana neighborhood, located next to the Pavão-Pavãozinho communities, all of which are equally dense in terms of population. The slum communities located up the hill from Copacabana have a “medium” HDI (0.64), while the wealthier neighborhood at the bottom of the hill had a “very high” HDI (0.93).

The high population density of both territories is an important factor in the virus’s rate of spread. The main difference pointed out by the simulator is that areas with less protection from Covid-19 (such as the Pavão-Pavãozinho communities) have more pronounced infection curves and therefore needed to isolate themselves more. In Figure 4, our simulations with 80% of the population practicing social isolation (a lockdown scenario) predicted a controlled epidemic curve on average for the rich part of the neighborhood (right) but not in the poorer communities (left).

**Figure 4:**
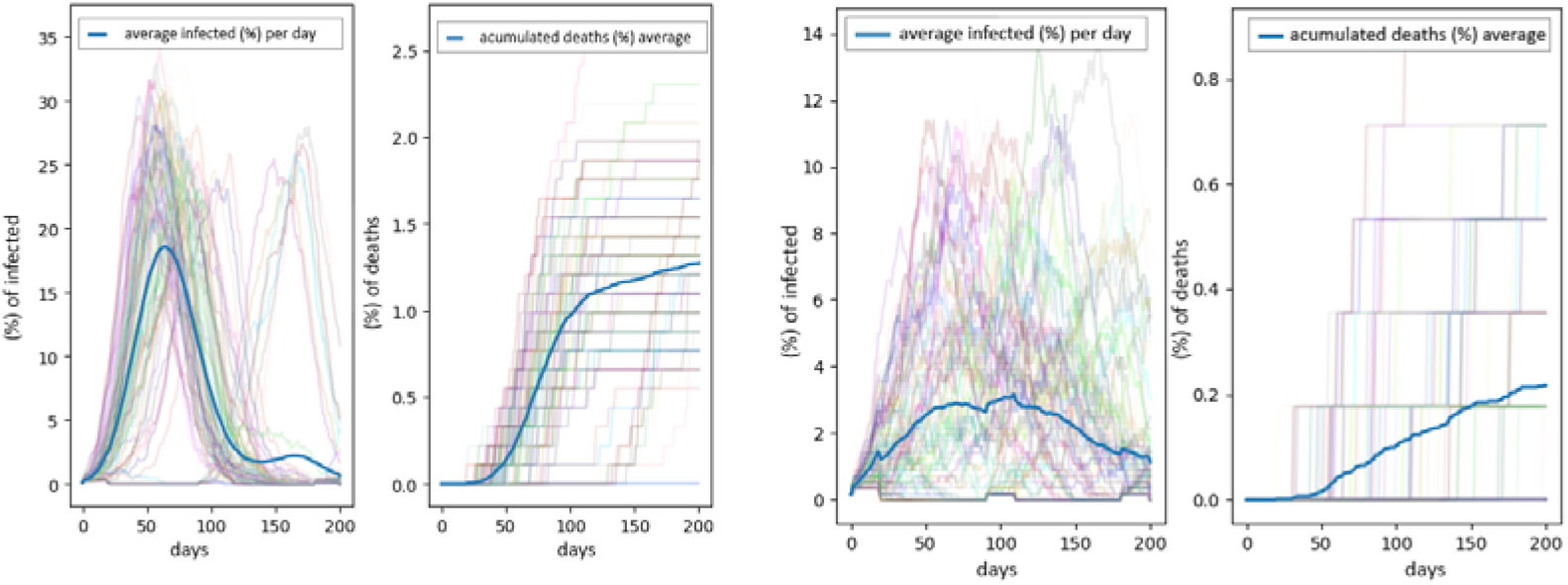
MD Corona simulations for Copacabana (right) and Pavão-Pavãozinho communities (left) with 80% social isolation. The colored lines represent each of the 100 simulations, and the blue line shows the average.

Similar simulations were made to compare the dynamics of the pandemic in poor and rich districts in the cities of Fortaleza (Meireles and Barra do Ceará, Ação Covid-19, 2020b), São Paulo (Brasilândia, Sapopemba, and Jardim Paulista, Ação Covid-19, 2020c), and Curitiba (Água Verde, Sítio Cerrado, and Tatuquara, Ação Covid-19, 2020e), and all showed similar results. What the model highlights is a new layer of inequality emerging in those cities as a result of unequal protection capacities across the cities’ regions.

One of the conclusions of these studies is that the government should urgently transfer resources intended for the richest districts and apply them to the poorest in order to mitigate the deadly effects of the pandemic.

### 4.2 A measurement of the effectiveness of NPI in Rio de Janeiro slums

In collaboration with the Observatorio de Favelas NGO, we developed a study (TEIXEIRA et al., 2020) to forecast the spread of the pandemic in rich and poor neighborhoods in different regions of the city of Rio de Janeiro. In this study, we were able to measure the extent to which the more organized communities of Maré and Rocinha worked to protect themselves from the virus when compared to other communities that were less organized, such as Cidade de Deus.

Given the initial conditions of vulnerability expressed in the CPI index map (Figure 5), Maré, Rocinha and Cidade de Deus have, respectively, ‘very low,’ ‘low’ and ‘medium’ protection indexes. Which, combined with the density of these territories (about 48,200, 30,400 and 28,700 inhab/km^2^, respectively), we would expect to lead to higher infection and death rates for the first two than the latter, which ended up not happening. Instead, based on a seroprevalence survey carried out in some communities within the city (Prefeitura da Cidade de Rio Janeiro, 2020) in late June, we know that the immunity rates in Rocinha and Maré were, respectively, 23% and 19%, whereas in Cidade de Deus it was 28%. After calibrating MD Corona for Cidade de Deus, we showed that by modifying the CPI index to “high” for Rocinha (from “very low”) and Maré (from “low”), the simulation yields, respectively, 25.7% and 21% of the population infected by the virus (see Figure 6 for Maré). Those results are compatible with the survey data for the same time frame, which shows the strength of the Model’s predictions.

**Figure 5:**
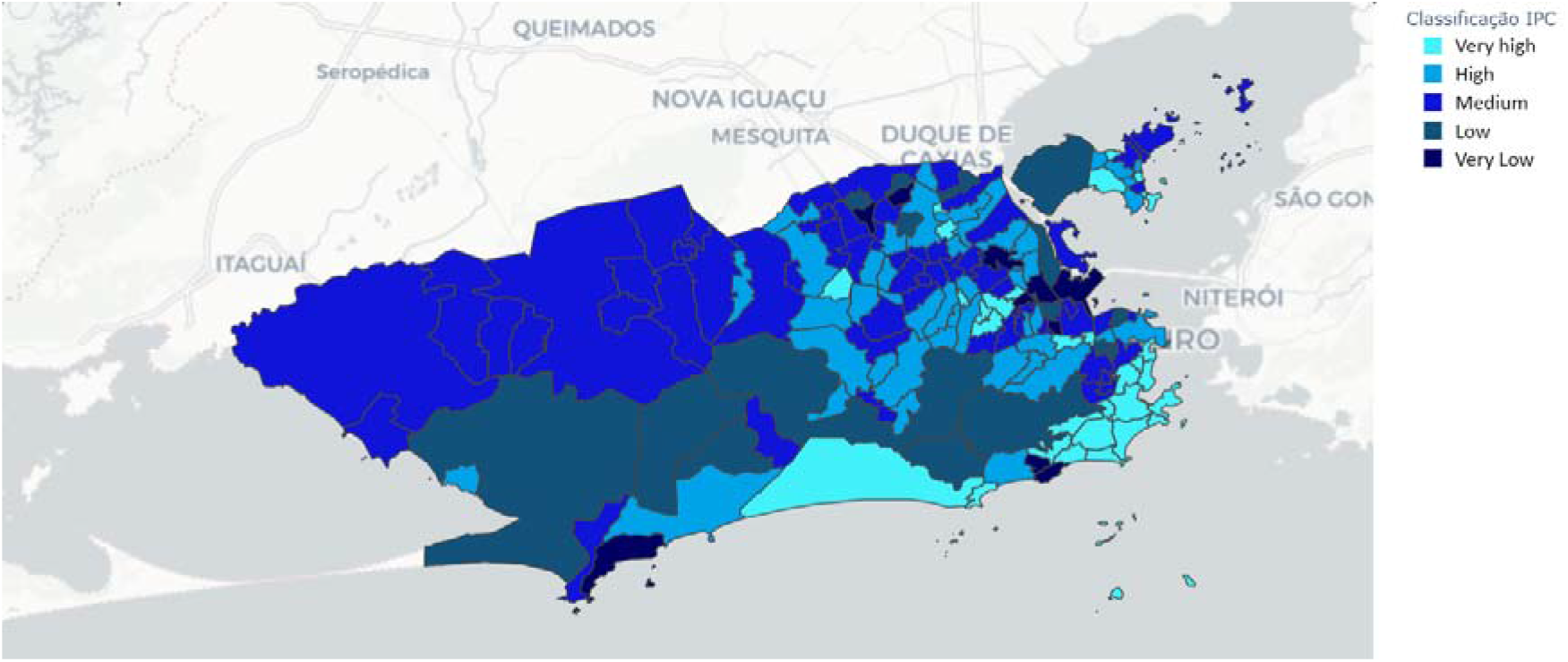
Coronavirus Protection Index (CPI) map (Ação Covid-19, 2020a) for the city of Rio de Janeiro.

**Figure 6:**
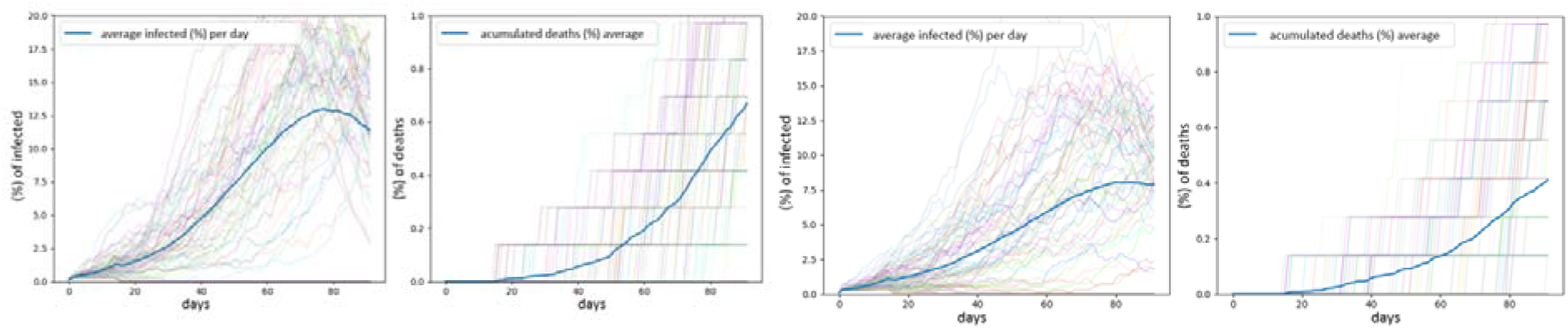
Simulation results for the Maré community with “low” CPI (left) - as indicated in our map in Fig. 5, and (right) with CPI increased to “high” after considering the effect of local actions. The colored lines represent each of the 100 simulations and the blue line shows the average.

One conclusion of this study is that actions implemented by Maré and Rocinha to contain the pandemic ended up raising the Coronavirus Protection Index (CPI) of these communities almost to the exact same CPI level as wealthier neighborhoods, such as Tijuca and Botafogo.

### 4.3 Local protection bubbles as an alternative explanation to herd immunity

More recently, we were faced with apparently contradictory results from the simulations that ended up demonstrating the robustness of the model. Seeking to alert public authorities to the possible dangers of reopening the economy in the city of São Paulo, we tried to model what would happen if social-isolation rates fell in the city of São Paulo starting in mid-July. We expected the infection curves to rise, but the model showed us the opposite: that even with the reduction in social isolation at that specific moment and in that specific territory, the curves fell, as shown in Figure 7.

**Figure 7.**
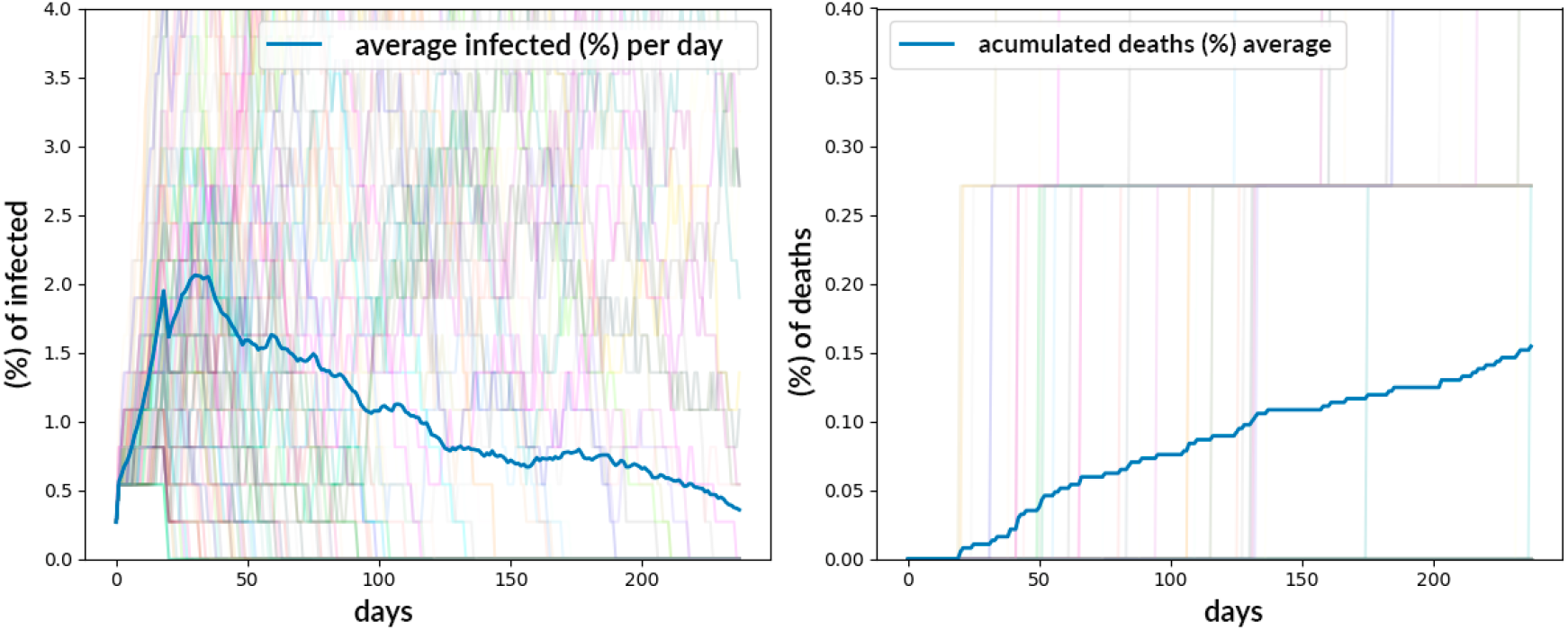
Result of MD Corona simulation based on trajectory of social distancing in the city of São Paulo, extended by 100 days with a 20% social-isolation rate.

At that time, scientists from different fields, including health care, estimated that the city could be close to so-called collective (or “herd”) immunity, or that this stage of immunity would require much lower rates than the 70% rate of infected as is usually expected.

Our simulations, however, showed that a more realistic explanation was the formation of “protection bubbles,” as highlighted in Figure 8, where a concentration of red (infected) agents surrounded by gray (immune) ones protects the susceptible agents (green) from being infected. In other words, social groups in which the infected dominate (with many already becoming immune) tend to have little contact with susceptible groups. Social isolation practices, whether voluntary or mandatory, can be part of the explanation for the origin of these bubbles, as well as socio-economic segmentation, which restricts contacts within the city, in addition to the new habits of the population (wearing masks, hand washing, etc.) that ended up shrinking the infection networks.

**Figure 8.**
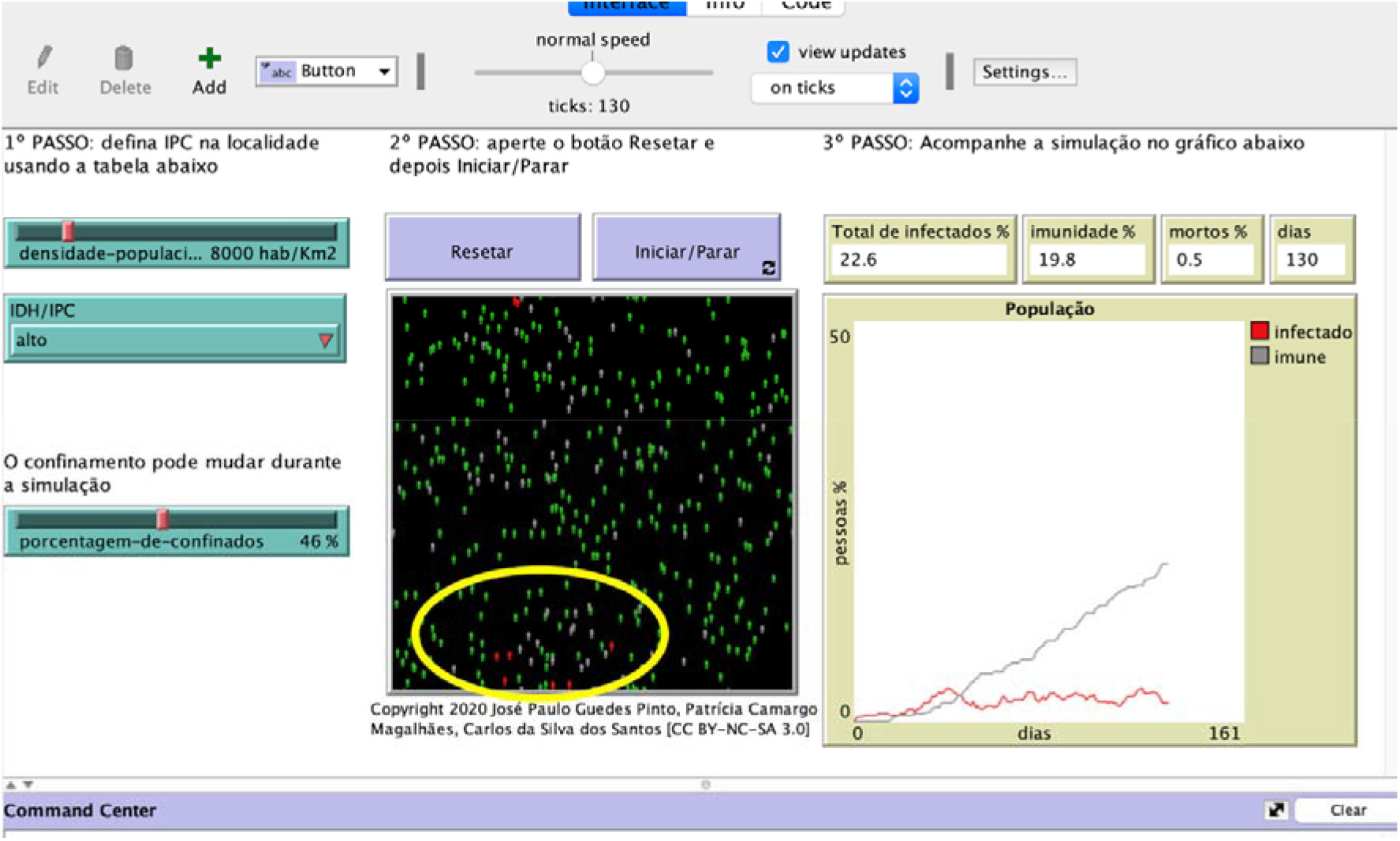
Grid from MD Corona simulation for the city of São Paulo. The yellow circle highlights the existence of protection bubbles.

We should note, however, that our hypothesis refers to a local equilibrium, possibly unstable, which can be perturbed by the introduction of a new infected agent, bursting these protection bubbles and restarting the infection networks. This study was published on the MedRxiv platform (Guedes Pinto et al, 2020) and also submitted to the PLOS One journal, and was widely discussed in the media as of the date of this article.

## 5. Conclusions

Modeling the pandemic has been tremendously important when alerting society to the importance of good decisions, changing the social culture, and improving scientific understanding of coronavirus dynamics, all of which help avoid an even worse tragedy.

In this work we presented MD Corona, a multiagent-model approach that predicts the coronavirus’s spread across different territories in unequal societies. The power of this model is that it is both intuitive and user-friendly, with good accuracy in its predictions. Although we calibrated the model using seroprevalence surveys, it does not depend on previous data to make predictions, as do the SIRS and statistical models.

Another important strength of our model is its ability to investigate very different scenarios by toggling only three parameters (population density, HDI/CPI indexes, and social isolation). It is also possible to change the other four parameters, including epidemiological ones, in the program code (available on our website) and adjusting them as the scientific literature on the pandemic advances.

MD Corona addresses different locations’ vulnerability to the spread of the virus by connecting the Coronavirus Protection Index – calculated based on social, health, and territorial infrastructure aspects – to the effective probability of virus transmission. In the context of an interdisciplinary research group, we simulated the virus’s epidemic curve in several major Brazilian urban centers. The results showed that areas with less protection from Covid-19 show more dramatic infection curves and require higher levels of social isolation. Those studies highlight the inequality of the initial conditions that locations may face in fighting Covid-19 and the existence of multiple epidemic curves within the same city.

The dynamics of the pandemic, however, change over time. An interesting outcome from one of our studies in the city of Rio de Janeiro showed that the NPI practiced by the Maré and Rocinha communities to contain the pandemic ended up raising their Coronavirus Protection Index to the level of wealthier neighborhoods. This result shows the model’s flexibility, as it denotes at least two ways of looking at the same parameter, namely, the effective probability of transmission between agents. It can be set as an initial structural condition (given by the HDI/CPI indexes) or as a practical response of populations that can change over time depending on several factors, such as the adaptation of the population to the pandemic, the capacity for collective action, major changes in the overall culture, etc.

Another important result that emerged was the capacity of the model to explain, in a simple way, why in some cities the virus’s spread decreased despite the reduction in social-distancing policies. We showed that this phenomenon is the result of an unstable equilibrium promoted by “local protection bubbles,” together with the exhaustion of contagion networks.

Recently, following global discussions about reopening schools, we adapted the model to simulate education environments like high schools, universities, kindergartens, etc. (Ação Covid-19 f). Although the simulator is less precise here than when examining cities, the insights it provides help government authorities decide in a more humane way whether or not to reopen schools. We focused on the school community to adapt parameters in the model that are specific to the school environment and their adherence to sanitary rules.

It is important to stress that there are still several other possibilities that can be explored within the model, such as changing the average immunity time of the population (decreasing it from six months to three months, for example), increasing or decreasing the average effective transmission probability of individuals, and increasing the number of people initially infected or already immune (or reintroducing more infected agents during the simulation).

In our first version of the model, back in March 2020, our simulations showed that the virus could be perpetuated in the environment for a long period (one year or more), leading to recurrent outbreaks and new waves of infection. This would happen if the virus were reintroduced into the system by external agents, if the period of immunity to the virus was short, or if the isolation of the population was not significant. Since then we have significantly improved the model by focusing on local short-term dynamics. But our initial intuition was confirmed by the actual data. The virus does demonstrate a cyclical behavior, and we will explore that in a forthcoming analysis. The same holds for the number of initial immune agents in the system, which is relevant at this stage of the pandemic, with a considerable number of people already affected.

Another important improvement could come in the simulation-analysis methodology. To deal with the stochastic behavior of the simulations we presented the average curve from 100 scenarios. However, in our next studies we want to perform a more complete analysis, identifying behavioral-change thresholds. In order to do that, we must analyze each simulation individually and then classify the different groups’ behaviors.

The study examples we discussed in the previous section shows that our model is robust and has become a tool for understanding Covid-19 dynamics, with potential uses in both outreach and communication and in supporting effective actions to mitigate the spread of the virus. The model’s potentialities and its applicability in the different scenarios described are possible because the model has been developed in the context of an interdisciplinary group – Ação Covid-19 – that seeks to use it as a tool to investigate the multiple faces of this unequal pandemic in Brazilian cities.

## Data Availability

Although this research used some literature data as inputs to the model, such as the incubation period, which is human, data from clinical or laboratory exams of patients were not used. Therefore, it is exempt from the appraisal of a research ethics committee by the Brazilian legislation expressed in the Resolution of the National Health Council (CONEP) number 466/12

## Acknowledgements

We would like to thank all members of the Ação Covid-19 group for fruitful discussions. This work was supported by Tide Setubal Foundation and the Federal University of ABC. PCM work was supported by Marie Curie (MSCA) grant no. 799974.

## Footnotes

1 They combine the description of different epidemiologic states through a SIR-based model with the commuting displacement and epidemic propagation through an agent-based approach. Although this model was very welcome in the science community for the ability to make predictions of the spread of the virus between cities in Sao Paulo State, the source and the simulator are not public.

2 It is the set of all parcels that are orthogonally adjacent to the region of interest (Weisstein, 2020).

